# Saliva as a non-invasive sample for the detection of SARS-CoV-2: a systematic review

**DOI:** 10.1101/2020.05.09.20096354

**Authors:** Zohaib Khurshid, Sana Zohaib, Chaitanya Joshi, Syed Faraz Moin, Muhammad Sohail Zafar, David J. Speicher

**Affiliations:** Department of Prosthodontics and Dental Implantology, College of Dentistry, King Faisal University, Al-Ahsa, Kingdom of Saudi Arabia; Department of Biomedical Engineering, College of Engineering, King Faisal University, Al-Ahsa, Kingdom of Saudi Arabia; Institute of Dentistry, School of Medicine, Medical Sciences & Nutrition, University of Aberdeen, Aberdeen, UK; National Center for Proteomics, University of Karachi, Pakistan; Department of Restorative Dentistry, College of Dentistry, Taibah University, Madinah Al Munawwarah, Kingdom of Saudi Arabia; Department of Laboratory Medicine, St. Joseph’s Healthcare Hamilton, Ontario, Canada; M.G. DeGroote Institute for Infectious Disease Research, Department of Biochemistry and Biomedical Sciences, DeGroote School of Medicine, McMaster University, Hamilton, Ontario, Canada

**Keywords:** saliva, diagnostics, COVID-19, SARS-CoV-2

## Abstract

The accepted gold standard for diagnosing coronavirus disease (COVID-19) is the detection of severe acute respiratory syndrome coronavirus 2 (SARS-CoV-2) RNA from nasopharyngeal swabs (NPS). However, shortage of reagents has made NPS collection challenging, and alternative samples need to be explored. Due to its non-invasive nature, saliva has considerable diagnostic potential. Therefore, to guide diagnostic laboratories globally, we conducted a systematic review to determine the utility of saliva for the detection of SARS-CoV-2. A systematic search of major databases (PubMed, ISI Web of Science, Scopus, and Google Scholar) was performed to identify published studies in accordance with the Preferred Reporting Items for Systematic Reviews and Meta-Analyses (PRISMA) guidelines. There was a total of 10 publications that fit the criteria for review. Most studies collected drooled whole saliva from hospitalized patients or pipetted saliva from intubated patients. Saliva was positive in 31-92% of patients depending on the cohort and length of hospitalization. Viral loads in saliva are comparable to those in NPS and ranged from 9.9 × 10^2^ to 1.2 × 10^8^ copies/mL during the first week of symptoms and decrease over time. Saliva can be positive up to 20 days post-symptom onset with viral loads correlating with symptom severity and degree of tissue damage. Based on these findings, we made suggestions to guide the clinical laboratory and suggest the need for diagnostic accuracy studies for the detection of SARS-CoV-2 from saliva.

## Introduction

The coronavirus disease (COVID-19) pandemic has affected the entire world, especially the most vulnerable population (i.e. the elderly living in nursing homes). The high demand of nasopharyngeal swabs (NPS), and the low supply of laboratory reagents and test kits highlight the need for alternative methods to facilitate accurate universal screening of COVID-19. The aetiological agent of COVID-19 is the novel *Betacoronavirus*, subgenus *Sarbecovirus*, severe acute respiratory syndrome coronavirus 2 (SARS-CoV-2), which originated in Hubei Province, China (1, 2). SARS-CoV-2 is a large, roughly spherical, enveloped virus, with a non-segmented positive-sense strand RNA genome ~30 kb in length (3). Since the initial outbreak, clinical symptoms have ranged from mild to severe atypical pneumonia with the disease spreading through human-to-human transmission (1). With an estimated incubation period of 5.1 days, and less than 2.5% of individuals displaying symptoms within 2.2 days of exposure, asymptomatic spread is possible, especially in children and healthy adults (4). The community transmission of SARS-CoV-2, especially asymptomatic spread, can be detrimental to both acute care hospitals and community settings, such as nursing homes. Therefore, robust diagnostics algorithms for SARS-CoV-2 are essential to quarantine infected individuals to prevent the spread of disease.

The current gold standard to detect SARS-CoV-2 RNA is by reverse transcription real-time polymerase chain reaction (RT-rtPCR) in NPS. NPS is invasive to collect, and due to widespread universal testing and a huge strain on supply lines, alternative diagnostic algorithms are needed (5-7). Saliva has considerable diagnostic potential: it is non-invasive, abundant, easily collected, and representative of oral and systemic health (8). The use of saliva looks promising as SARS-CoV-2 RNA is present in saliva with loads and sensitivity comparable to NPS (9-11). However, salivary endonucleases make proper sample handling critical for accurate testing (12). Therefore, we conducted a systematic review to evaluate the potential of using saliva for the detection of SARS-CoV-2 and make suggestions for the clinical diagnostic labotatory.

## Methodology

### Literature Search and Selection Criteria

This scoping review follows the guidelines of Preferred Reporting Items for Systematic Reviews and Metanalysis (PRISMA). A systematic search was performed on four major databases (PubMed, ISI Web of Science, Scopus, and Google Scholar) by two independent reviewers to identify articles published in English prior to April 25, 2020 to answer, “Can saliva be used for to detect SARS-CoV-2 and diagnose COVID-19?” The systematic search was performed using the following Medical Subject Headings (MeSH) terms and keywords: “COVID-19” OR “COVID-2019” OR “severe acute respiratory syndrome coronavirus 2” OR “2019-nCoV” OR “SARS-CoV-2” AND “saliva”. Manuscripts were included if they aligned with the following PECOS (Patient, Exposure, Comparator and Outcome) guidelines: (P) male or female of any age group visiting the Emergency Department; (E) SARS-CoV-2 infected; (C) Systematically healthy patients as control; (O) salivary viral load. Manuscripts were excluded if they were letters to the editor, perspectives or review papers. Given the explosion of new research, preprints were included from BioRxiv and medRxiv if the inclusion criteria were met. Both reviewers mutually agreed to use the same inclusion and exclusion criteria for the search, and disagreements were resolved by discussion. Ethics approval was not required.

### Data Extraction

Data extraction was performed independently by two investigators and confirmed by a third. Each study was examined for saliva sampling protocol, nucleic acid extraction procedure, SARS-CoV-2 detection, and primary findings. Discrepancies were resolved by consensus.

## Results

### Search Results and Cohorts Studied

We identified 25 potential articles from database searches that investigated the detection of SARS-CoV-2 in saliva (Figure 1). After removing the duplicates and reviewing the abstracts for inclusion and exclusion criteria, the full-text of 10 papers were scrutinized for this systematic review. Studies were from China (n=10), Italy (n=2), USA (n=1) and South Korea (n=1). Apart from case reports, studies enrolled hospitalized patients and compared saliva to NPS for the initial diagnosis and viral load monitoring. Apart for two studies, study cohorts were small (range: 12-44) with a relatively equal number of males and females with an average age of 61 years (range: 18-92 years) (Table 1). The specimen collected varied by study and included saliva (n=6), sputum or deep saliva (n=4), and oropharyngeal swabs (OPS, n=3). No study revealed transportation or storage conditions. There is great variation in the extraction and amplification kits used, but most extractions were performed manually with commercial spin columns (n=6), and two studies did automated extraction with the NucliSENS^®^ easyMag^®^ (9, 13). PCR amplification was performed with a mix of lab-developed tests (LDT, n=6) and commercial assays (n=4) targeting a wide range of genes.

**Figure 1:**
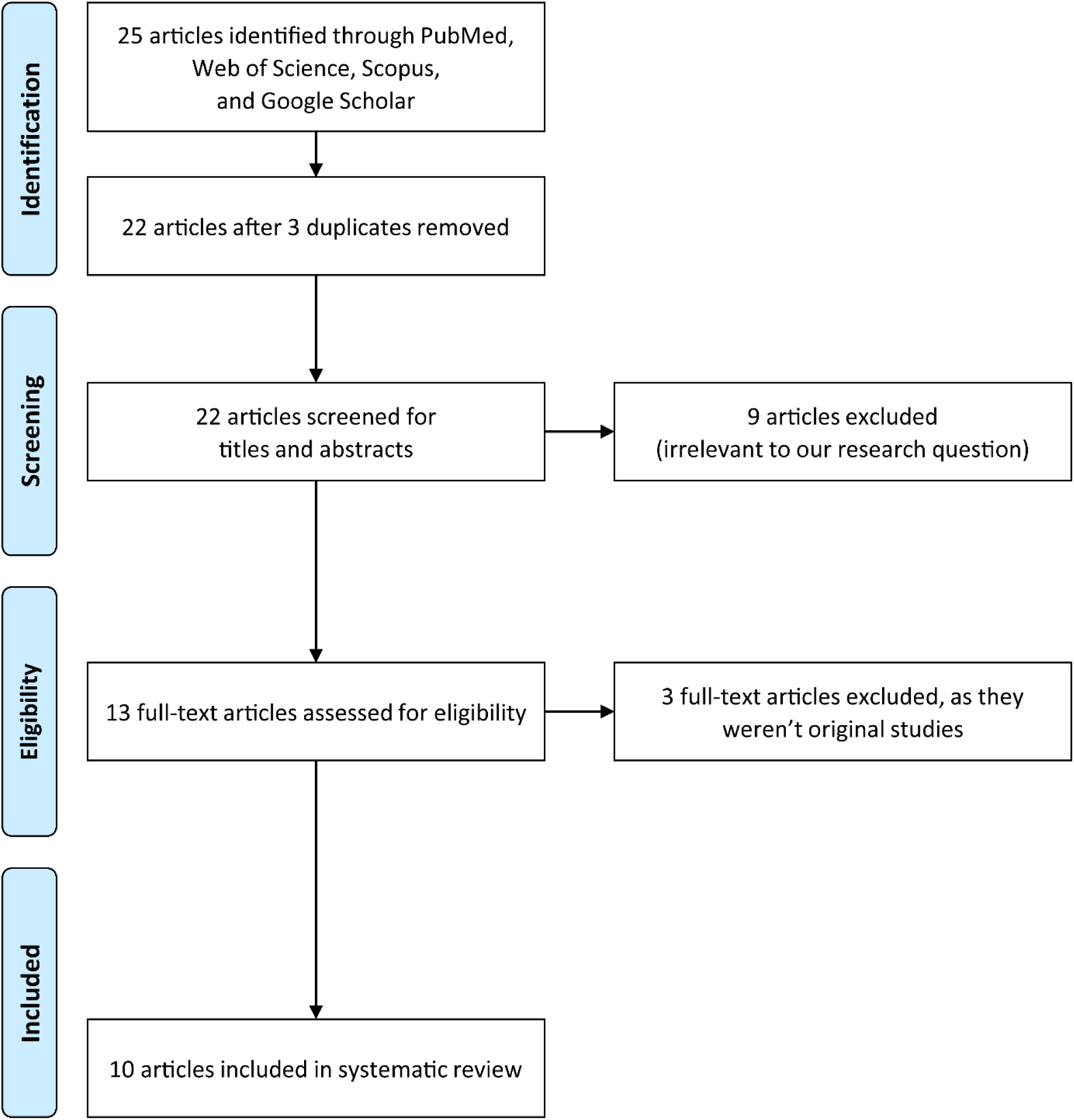
A PRISMA flow diagram of the search strategy for saliva, COVID-19, and SARS-CoV-2.

**Figure 2:**
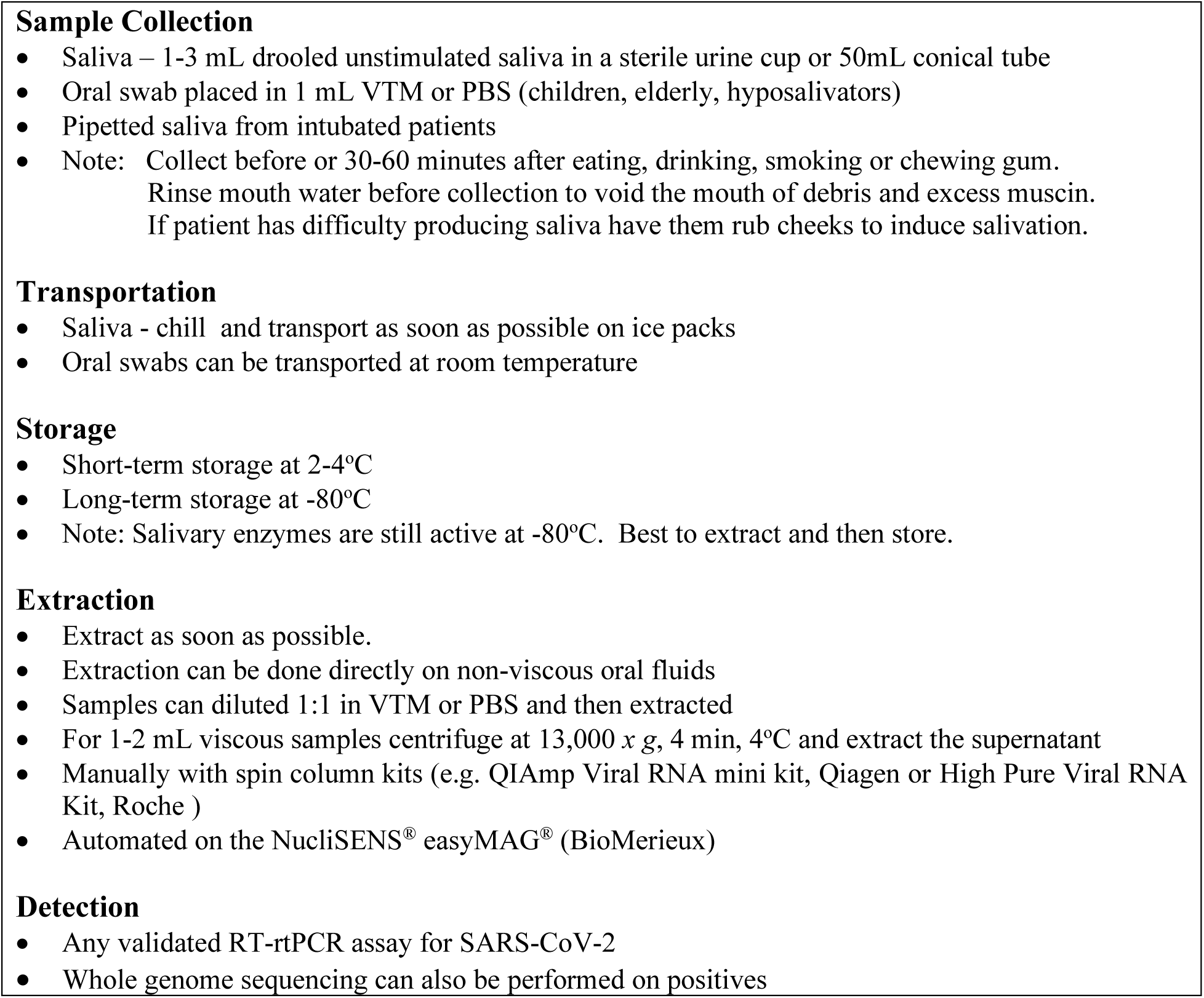
Suggested method for collection and processing of saliva for the detection of SARS-CoV-2.

**Table 1:** Systemic search findings evaluating saliva for the detection of SARS-CoV-2 RNA.

| S/N | Author, year;<br>(Country) | Study participants | Specimen Collected | Sample extraction and<br>Diagnostic Assay | Summary of Results | Reference |
| --- | --- | --- | --- | --- | --- | --- |
| 1 | Azzi et al., 2020<br>(Italy) | 25 COVID-19 positive patients with severe or very severe disease<br>17 males, 8 females<br>mean age 61.5 years<br>(range: 39 to 85 years) | NPS<br>Drooled saliva<br>Pipetted saliva if intubated<br>2<sup>nd</sup> oral sample 4 days later | Saliva resuspended in 2mL PBS<br>RNA extraction with QIAmp Viral RNA mini kit (Qiagen, Germany)<br>One step RT-rtPCR using Luna Universal qPCR Master Mix (New England BioLabs Ltd., USA)<br>RNA Target: 5'UTR | First saliva sample from all patients was positive (Ct values: mean 27.2, range 18.1- 32.2).<br>Inverse correlation between LDH values and the Ct values in saliva shows that viral load in saliva correlates to tissue damage<br>Second saliva sample from 8 patients positive<br>2 patients were saliva positive salivary but NPS was negative on the same day | (17) |
| 2 | Azzi et al., 2020<br>(Italy) | 2 COVID-19 positive men (64 & 71 years) | NPS<br>BAL<br>Drooled saliva | Saliva resuspended in 2mL PBS<br>RNA extraction with QIAmp Viral RNA mini kit (Qiagen, Germany)<br>One step RT-rtPCR using Luna Universal qPCR Master Mix (New England BioLab s Ltd., USA)<br>RNA Target: 5'UTR | In both patients, saliva was positive when both NPS and BAL were negative | (26) |
| 3 | Chen et al., 2020<br>(China) | 31 COVID-19 positives (15 males, 16 females)<br>median age of 60.6 years (range: 18 to 86 years).<br>5/31 were critically ill and on ventilator support. | 1.5mL midstream salivary fluid with cotton swabs<br>Oropharyngeal swabs | Extraction method not disclosed<br>RT-qPCR using a commercial test kit (BioGerm. InC, China) on a Roche Cobas z480 PCR Analyzer<br>RNA Target: ORF1ab and N | 13/31 (42%) positive in OPS.<br>4/13 (31%) OPS positives were saliva positive<br>3/4 (75%) of saliva positive patients were critically ill and on ventilators | (27) |
| 4 | Cheng et al., 2020<br>(Hong Kong, China) | 42 COVID-19 positive patients (20 males, 22 females) | Upper respiratory specimens (i.e. NPA, flocked swabs, and throat swabs)<br>Lower respiratory | Total nucleic acid (TNA) extraction with NucliSENS® easyMAG® (BioMerieux, Canada). | The viral loads of the first confirmed case were $3.3 \times 10^6$ copies/mL in the pooled nasopharyngeal and throat swabs, whereas $5.9 \times 10^6$ copies/mL in saliva on the same day. | (13) |
|  |  | median age of 59 years<br>(range, 22–91 years) | specimens (i.e. sputum,<br>endotracheal aspirates, or<br>BAL) | RT-PCR using the LightMix<br>Modular SARS and Wuhan CoV E-gene mix (TIB Molbiol, Germany) |  |  |
| 5 | Han et al., 2020<br>(South Korea) | 27-day old neonate and her<br>mother diagnosed with<br>COVID-19 | NPS, OPS, Stool, Saliva,<br>Plasma, and Urine | RNA extraction using MagNA Pure<br>96 DNA and Viral NA small<br>volume kit (Roche, Germany)<br>RNA detected using the<br>PowerChek™ 2019-nCoV Real-time PCR Kit (Kogene Biotech,<br>South Korea)<br>RNA Target: ORF1b and E | • In neonate, SARS-CoV-2 RNA was detected in all clinical specimens<br>• Saliva was positive in neonate but not in mother<br>• Neonate's saliva sample was positive up to 10 days after symptoms onset.<br>• Viral load in neonate's stool and urine remained high up to 18 days even after the respiratory specimens became negative<br>• Mother's plasma, urine, and breast milk tested negative | (28) |
| 6 | To et al., 2020<br>(Hong Kong, China) | 12 COVID-19 positive patients<br>(7 males, 5 females)<br>median age: 62.5 years<br>(range: 37–75 years) | NPS<br>Sputum (cough out saliva<br>from throat into a sterile<br>container) collected 0–7 days<br>post hospitalization | Total nucleic acid (TNA) extraction<br>with NucliSENS® easyMag®<br>(BioMerieux, Canada)<br>In-house 1-step RT-qPCR<br>Assay using QuantiNova SYBR<br>Green RT-PCR Kit (Qiagen,<br>Germany) in a LightCycler 480<br>Real-Time PCR System (Roche,<br>Germany)<br>RNA Target: S gene | • 11/12 (91.7%) patients were saliva positive<br>• Median viral load in saliva $3.3 \times 10^6$ copies/mL (range, $9.9 \times 10^2$ to $1.2 \times 10^8$ copies/mL)<br>• Viral load highest in earliest available specimen for 5 patients (83.3%)<br>• In 1 patient saliva was positive 11 days post-hospitalization<br>• In 33 patients whose NPS tested negative corresponding saliva specimens also tested negative | (9) |
| 7 | To et al., 2020<br>(Hong Kong, China) | 23 patients<br>(13 males, 10 females),<br>median age 62 years<br>(range: 37–75)<br>10 patients (43.5%) had<br>severe COVID-19 requiring<br>oxygen supplementation.<br>13 patients had mild disease. | Blood, Urine, Posterior<br>oropharyngeal saliva, and<br>Rectal swabs<br>Saliva was collected early<br>morning from the<br>posterior oropharynx (ie,<br>coughed up by clearing<br>the throat) before<br>toothbrushing and breakfast. | Extraction method not disclosed<br>in-house RT-qPCR targeting the<br>SARS-CoV-2 RNA-dependent-RNA-polymerase-helicase gene<br>region | • Viral load in posterior oropharyngeal saliva samples was highest during the first week of symptom onset then gradually declined.<br>• 3/23 (13%) patients did not produce a positive saliva sample<br>• 7/21 (33%) patients had viral RNA detected in saliva sample for 20 days or longer after the onset of symptoms.<br>• No association between prolonged detection of viral RNA and severity of illness. | (14) |
|  |  |  | If patients were intubated, an endotracheal aspirate was obtained. |  |  |  |
| 8 | Wyllie et al., 2020 (USA) | 44 COVID-19 positive (23 males, 21 females), mean age 61 years (range 23–92)<br>10 (23%) required mechanical ventilation, and 2 (5%) died.<br>98 asymptomatic healthcare workers were enrolled | NPS and saliva was collected every 3 days.<br>Saliva was self-collected by the patient first thing in the morning prior to food or brushing of teeth. Saliva collected by spitting into a sterile urine cup until ~1/3 full (excluding bubbles) | Total nucleic acid extracted from 300µl of viral transport media from NPS or 300µl of whole saliva using MagMAX Viral/Pathogen Nucleic Acid Isolation kit (ThermoFisher Scientific, USA).<br>US CDC RT-rtPCR primer/probe sets for 2019-nCoV_N1 and 2019-nCoV_N2 and the human RNase P (RP) as an extraction control. | • Viral loads 5X higher in saliva than in NPS<br>• 21% saliva positives did not have a matched NPS positive.<br>• 8% NPS positives did not have a matched saliva positive.<br>• Saliva yielded greater overall sensitivity.<br>• 2 asymptomatic HCW were saliva positive NPS-negative. | (15) |
| 9 | Zheng et al., 2020 (China) | 96 consecutively admitted patients with laboratory confirmed SARS-CoV-2 infection: 22 with mild disease and 74 with severe disease. Median age was 55 years (range: 44.3-64.8). | Respiratory, Serum, Stool, and Urine samples were collected daily.<br>Sputum collected from the respiratory tract of patients with sputum. In patients without sputum saliva was collected after deep cough. | Viral RNA was extracted using the MagNA Pure 96 (Roche, Germany).<br>RT-qPCR was performed using a China Food and Drug Administration approved commercial kit specific for SARS-CoV-2 detection (BoJie, China).<br>RNA Target: ORF1ab | • Infection was confirmed in all patients by testing sputum and saliva samples.<br>• Higher viral loads were found in patients with severe disease compared to those with mild disease. | (16) |
| 10 | Zhang et al. 2020 (China) | 178 COVID-19 positive hospitalized patients | Oral swabs, anal swabs and blood samples.<br>For swabs, 1.5 ml DMEM+2% FBS medium was added in each tube. Supernatant was collected after 2500 rpm, 60 s vortex and 15–30 min standing. | RNA was extracted from 200 µL of samples with the High Pure Viral RNA Kit (Roche, Germany).<br>qPCR by HiScript® II One Step RT-qPCR SYBR® Green Kit (Vazyme Biotech Co., Ltd, China) in an ABI 7500.<br>RNA Target: S gene | • In the first study, after days of medical treatments 53.3% were oral swabs positive, 27% were anal swabs positive, and 40% were serum positive.<br>• In the second study, at day 0, 80% of positives came from oral swabs, but on day 5, 75% anal swabs and 50% oral swabs were positive. Possible shift in virus location as disease progresses. | (18) |

NPS=nasopharyngeal swabs, OPS=oropharyngeal swabs, BAL=bronchoalveolar lavage, RT-PCR=reverse transcription polymerase chain reaction, RT-qPCR=quantitative reverse transcription polymerase chain reaction

### Salivary Diagnostics

Despite the heterogeneity of oral samples used, based on these limited studies, it is evident that saliva is comparable, if not superior to NPS for initial detection of SARS-CoV-2 upon hospitalization and is more consistent for monitoring viremia. In most comparative studies, drooled saliva had higher positivity rates than deep saliva/sputum and was useful from initial screening to intubation. Positivity of saliva was 31-92% depending on the cohort and length of hospitalization. Viral load in saliva was highest during the first week of symptom onset, ranging from 9.9 × 10^2^ to 1.2 × 10^8^ copies/mL, and then gradually declined (9, 14). At initial screening viral loads and positivity rates were comparable to NPS, but one study found salivary loads were five-times higher than in NPS (15). Higher salivary viral loads were also found in patients with more severe disease (16) and correlated to tissue damage (17). In a study screening 98 asymptomatic health care workers all NPS were negative, but two were positive in saliva (15). Studies monitoring viral loads reported that initially saliva was comparable to NPS, but over time NPS became negative while saliva remained positive up to 20 days post-symptom onset, even after respiratory symptoms became negative. However, after the first week positivity rates in saliva decreased, and positivity in anal swabs increased, suggesting a possible shift in viral infection as disease progresses (18).

## Discussion

Initial studies examining the utility of saliva for the detection of SARS-CoV-2 RNA are comparable, if not superior to NPS for screening and monitoring viral loads. With NPS reagents increasingly in short supply, saliva is a possible alternative sample for diagnostic algorithms. Infectious cell-free SARS-CoV-2 virion is transmitted in salivary droplets by infected people breathing, talking, coughing, or sneezing in close contact and infecting another nearby person through the mouth, nose or eyes (10). SARS-CoV-2 infects human epithelial cells through the host cell receptor angiotensin-converting enzyme II (ACE2), which is expressed on cells lining the lungs, oral buccal and gingiva (19). This infectious virion is both detectable in saliva and culturable on Vero E6 cells making saliva both a non-invasive sample that is easy to collect but also a potential exposure risk for front-line healthcare workers (9).

SARS-CoV-2 has been detected in a variety of oral samples, including whole saliva, oral swabs, oropharyngeal swabs, and deep saliva/sputum. There is a wide range of commercially available saliva collection devices but given the high sampling demand and limited stocks of swabs, 1-3 mL of unstimulated whole saliva expectorated into a sterile urine container or 50 mL conical tube is the easiest and most reliable sample to collect, unless the patient is a hyposalivator or intubated. For an intubated patient, young child, or elderly person pipetted saliva or oral swab placed in 1 mL viral transport media (VTM) will suffice (17, 20). Saliva should always be collected before or 30-60 minutes after eating, drinking, smoking or chewing gum with the mouth rinsed with water prior to collection to void the mouth of debris (21). If a patient is having difficulty salivating rubbing the outside of the cheek may help. As sample positivity is essential in hospitalized patients, due to the effect of diurnal variation on levels of salivary biomarkers, two studies collected saliva first thing in the morning prior to food or brushing of teeth (14, 15). The other studies did not account for circadian rhythm, but found the viral load to be highest in saliva for the first week after the onset of symptoms, suggesting that adequate salivary samples can be collected at anytime. However, due to salivary enzymes, it is essential, especially if samples require transportation, that whole saliva be chilled immediately after collection (e.g. shipped on ice packs) or stored in a commercial stabilizer (e.g. RNAlater and VTM) and extracted as soon as possible (12, 22).

Extraction of SARS-CoV-2 RNA from saliva samples has been done with commercial spin columns or automated on the NucliSENS^®^ easyMAG^®^. Processing saliva can be laborious as viscosity varies greatly and samples high in mucin can clog spin columns and automated extractors. To overcome this problem, several studies mixed saliva in 2 mL VTM or PBS prior to extraction. As saliva contains cell-free infectious virion, it may be possible to centrifuge the saliva collected in sterile 50 mL conical tubes at 2,800 *x* g,10-minutes, 4°C, aspirate the supernatant, and proceed with extraction (22). Small samples (1-2 mL) can be centrifuged at 13,000 *x* g, 4 minutes, 4°C in microcentrifuge tubes. Detection of SARS-CoV-2 RNA can then be performed with any validated LDT or commercially available assay (5). Subsequent whole genome analysis on positives using Oxford Nanopore MinION or by bait capture hybridization probes coupled with Illumina sequencing for surveillance and outbreak analysis (14, 23).

The utility of saliva for the detection of SARS-CoV-2 appears clinically useful, but controlled diagnostic accuracy studies comparing saliva to matched NPS in positive and negative patients are desperately needed. As outlined by the US FDA, the use of saliva for the detection of SARS-CoV-2 requires a clinical study comparing whole saliva to NPS collected at the same time from a minimum of 30 positive/30 negative pairs with discordant results resolved with further testing (24). Similar validations could also be performed for other types oral fluids. Future studies could also compare salivary and serum IgG as a screen for immunity (25). Nonetheless, properly conducted comparative studies must be performed as soon as possible to help direct the clinical laboratory in the fight against COVID-19.

## Conclusion

In conclusion, saliva can be used to detect SARS-CoV-2 in both symptomatic patients and asymptomatic carriers. Studies to date have shown that viral loads in saliva are comparable or higher than in NPS after the onset of symptoms and remain detectable in saliva after respiratory symptoms dissipate and NPS test negative. However, due to salivary enzymes, samples must be handled correctly and processed in a timely manner. Well conducted diagnostic accuracy studies are desperately needed to validate the use of saliva for the diagnosis of COVID-19.

## Data Availability

All data is publically available.

